# Prevalence and pattern of refractive errors among Yanomami Indigenous people in the Brazilian Amazon: a cross-sectional observational study

**DOI:** 10.64898/2026.05.25.26354064

**Authors:** Maria Christina Chagas Ferreira, Marcos Antonio Pellegrini, Bianca Jorge Sequeira

## Abstract

**Background:** Refractive errors are the leading cause of preventable visual impairment worldwide, yet data from isolated Indigenous populations remain virtually absent from the global literature. The Yanomami, one of the largest Indigenous peoples in the Americas with recent and limited contact with non-Indigenous society, have no prior epidemiological data on refractive errors.

**Methods:** A cross-sectional observational study was conducted in 2024 at the Yanomami Indigenous Health House, Boa Vista, Roraima, Brazil. A total of 158 self-identified Yanomami individuals aged 5 years or older were examined by an ophthalmologist. Refractive status was classified according to International Myopia Institute criteria.

**Results:** Emmetropia was observed in 67.7% of participants, with a marked age-related decline from 100% in children aged 5 to 9 years to 38.6% in those aged 40 to 59 years. Myopia was present in 16.5% of participants, all low myopia; it was absent in children under 10 years and no high myopia was identified. Astigmatism affected 24.1% of participants and hyperopia 13.3%. Presbyopia was identified in 25.9%. Overall, 25.3% of participants presented with reduced visual acuity attributable to uncorrected refractive error, of whom 67.5% improved to normal or near-normal acuity (p < 0.001).

**Conclusions:** This is the first characterisation of the Yanomami refractive profile, revealing a distinct myopia pattern shaped by high outdoor exposure and minimal near-work demands. Despite this, refractive correction remains effectively inaccessible to this population, leaving preventable visual impairment unaddressed and reflecting a profound health inequity. Corrective lens provision represents a high-impact, scalable intervention for this underserved community.

## Introduction

Refractive errors are the leading cause of visual impairment worldwide^1^ and one of the most preventable contributors to functional vision loss across all age groups.^2^ According to global estimates, uncorrected refractive error accounts for a substantial proportion of moderate and severe visual impairment, with significant consequences for education, productivity, social participation, and quality of life.^1^ Despite the simplicity and cost-effectiveness of refractive correction, access to vision assessment and corrective lenses remains highly unequal, disproportionately affecting populations living in remote, low-resource, and underserved settings.

Indigenous peoples experience persistent health inequities driven by geographic isolation, socioeconomic vulnerability, and limited access to culturally appropriate health services.□ In the Americas, these disparities are particularly pronounced in remote regions of the Amazon, where logistical barriers, workforce shortages, and linguistic diversity restrict the availability of specialized eye care.□As a result, visual impairment due to uncorrected refractive error often remains undiagnosed and untreated, even when effective interventions are readily available.□□ Evidence on refractive errors in Indigenous populations is scarce and heterogeneous.□□ Existing studies have largely focused on urban or semi-urban Indigenous groups or have addressed specific ocular conditions rather than population-level refractive profiles.□ Data from Indigenous peoples living in areas of recent contact with non-Indigenous society are especially limited, constraining the development of targeted public health strategies. Environmental, cultural, and lifestyle factors, including high levels of outdoor activity, limited formal education, and reduced exposure to prolonged near work, suggest that the distribution and pattern of refractive errors in these populations may differ from those observed in urban settings, underscoring the need for context-specific epidemiological data.

The Yanomami are one of the largest Indigenous groups in the Brazilian Amazon and are characterized by high territorial mobility, geographic isolation, and a history of relatively recent and limited contact with non-Indigenous society. Eye health data in this population are particularly scarce. While previous studies have documented a high burden of visual impairment and blindness among the Yanomami, uncorrected refractive error has consistently emerged as a leading and potentially avoidable cause of vision loss.^1^□ However, detailed information on the prevalence, type, and distribution of refractive errors in this population remains lacking.

This study aimed to estimate the prevalence and pattern of refractive errors among Yanomami Indigenous people living in the Brazilian Amazon and to assess the functional impact of refractive correction on visual acuity.

## Methods

### Study design and setting

This was a cross-sectional, observational study conducted between June and August 2024 at the Yanomami Indigenous Health House (Casa de Saude Indigena Yanomami, CASAI-Y), located in Boa Vista, Roraima, Brazil. CASAI-Y is a regional referral facility within the Yanomami Special Indigenous Health District (DSEI-Y), providing care to Indigenous individuals from remote communities across the Yanomami Indigenous Territory who require health services unavailable at primary care units. Owing to geographic, legal, and logistical constraints that preclude population-based surveys within the Yanomami Indigenous Territory, CASAI-Y represents the most feasible setting to access a diverse and demographically representative cross-section of this highly mobile Indigenous population.

The study included self-identified Yanomami Indigenous individuals aged 5 years or older who were present at CASAI-Y during the data collection period. Exclusion criteria included refusal to participate, inability to complete the ophthalmological examination due to cognitive impairment or insufficient clinical conditions, or refusal of any component of the evaluation.

Participation was voluntary and based on informed consent; for minors, consent was obtained from a parent or legal guardian, with assent from the child when appropriate. All informed consent and assent documents were translated into the participants’ native language and explained orally by a native-speaking Indigenous interpreter. Given the high prevalence of illiteracy among participants, the consent process was audio recorded to ensure full ethical compliance and participant comprehension. In cases in which written consent could not be provided, the interpreter signed the consent form on the participant’s behalf, in accordance with the approved protocol.

### Study population and sample

At the time of the study, 278 eligible individuals aged 5 years or older, belonging to four officially registered Yanomami subgroups (Yanomami, Sanuma, Xirixana, and Xiriana), were present at the facility. A total of 158 individuals met the inclusion criteria and agreed to participate, corresponding to 56.8% of the eligible population. The participant flow is presented in Figure 1.

**Figure 1.**
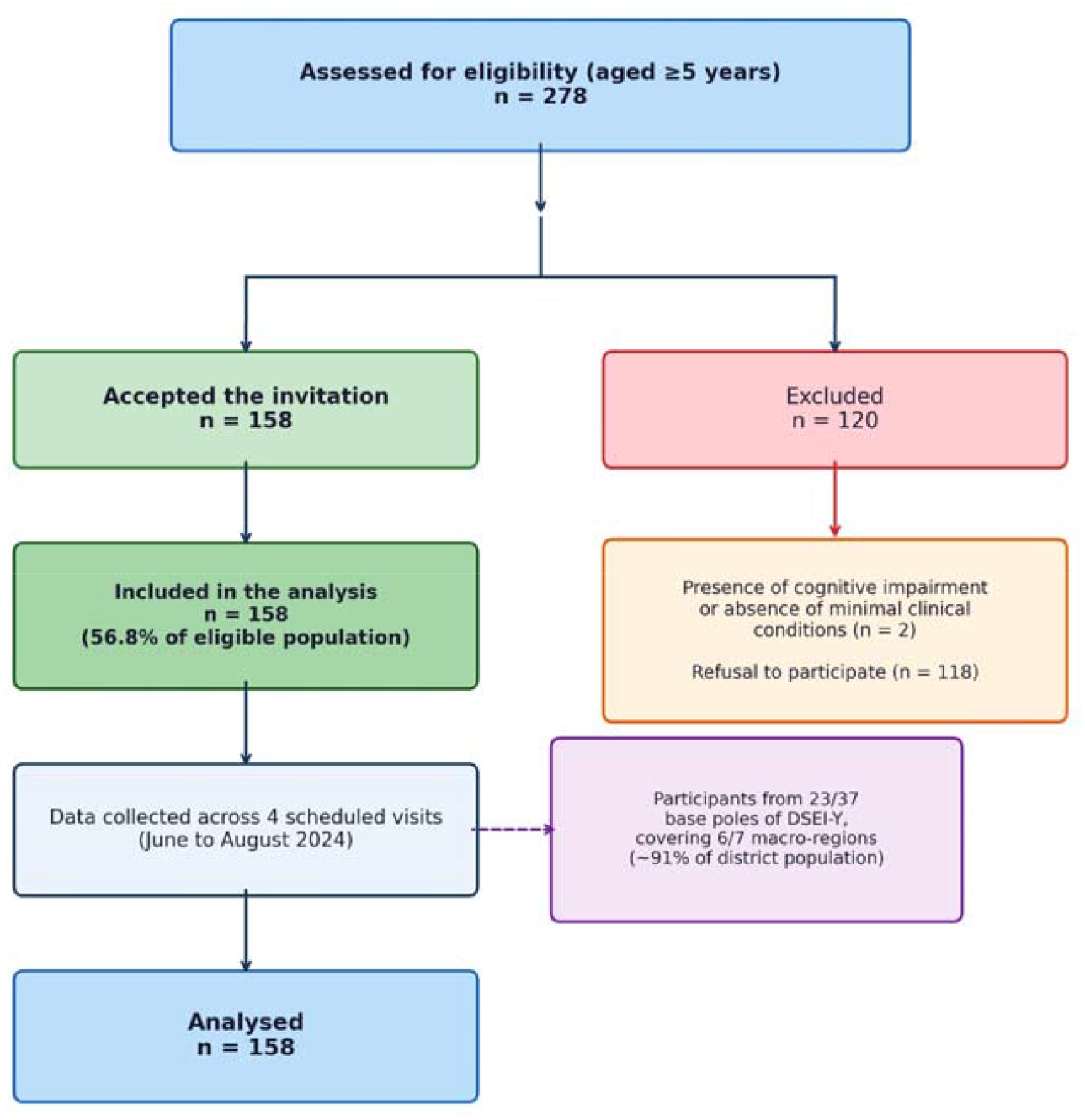
Flow diagram showing recruitment and inclusion of participants in the study (2024).

The sample size was not predefined by probabilistic sampling; rather, it was based on the total accessible population during the study period. Nevertheless, the achieved sample exceeded the minimum estimated size required to obtain a 95% confidence level with a 5% margin of error in a homogeneous population.

Participants originated from 23 of the 37 base poles of the DSEI-Y, covering six of the seven macro-regions of the Yanomami territory and representing approximately 91% of the district population. A moderate positive correlation (Pearson’s r = 0.57) was observed between the number of examined individuals per base pole and the total population size of each locality, suggesting proportional geographic representation. Given the non-random nature of recruitment and the facility-based design, potential selection bias cannot be excluded; however, the proportional geographic distribution observed supports partial representativeness of the broader DSEI-Y population.

### Data collection and ophthalmological examination

Data collection occurred during four scheduled field visits and was organised according to a structured, three-step ophthalmological assessment protocol. In the initial step, participants’ demographic information (sex, age, community of origin, and reference health post) was recorded, and informed consent or assent procedures were completed.

The second step consisted of objective ophthalmological measurements performed by trained personnel. Refractive screening was conducted using a Welch Allyn Spot Vision Screener handheld photoscreener. Intraocular pressure was measured with a Huvitz HNT-7000 non-contact tonometer, and retinal images were obtained using an Eyer portable fundus camera to document posterior segment findings and to rule out ocular conditions that could influence refractive assessment.

Immediately after the objective measurements, the third step involved a clinical interview to collect self-reported ocular history and general health information. This was followed by slit-lamp biomicroscopy performed with a portable slit lamp and subjective refraction carried out by an ophthalmologist.

Distance visual acuity was measured monocularly and without correction using a Snellen “E” chart, as recommended by the Brazilian Ministry of Health, and a culturally adapted pictorial chart specifically developed for the Yanomami population. Both charts were positioned at a distance of three meters from the participant, in an adequately illuminated environment. For participants with presenting visual acuity worse than 20/30 or who reported visual difficulties, best-corrected visual acuity was assessed dynamically using a Greens refractor.

Near visual acuity was evaluated in participants aged 35 years or older using a Jaeger near vision card with the “E” optotype and a corresponding pictorial version adapted for this study. Testing was performed at a standard distance of 37 cm, and near addition was determined using the Greens refractor when required.

Throughout all interviews and examinations, Indigenous interpreters fluent in the participant’s specific dialect facilitated communication. No adverse events associated with the use of cycloplegic agents were observed. At the end of the assessment, prescriptions for corrective lenses were issued when indicated, and participants requiring further care were referred through the Brazilian Unified Health System (SUS).

### Definitions and outcomes

Refractive status was classified according to the International Myopia Institute (IMI) standards.^1^□ Spherical equivalent (SE) was calculated for each eye. Myopia was defined as SE <= -0.50 D, hyperopia as SE >= +0.50 D, and astigmatism as absolute cylinder >= 1.00 D in at least one eye. Combined refractive errors were further categorized as compound myopic astigmatism, compound hyperopic astigmatism, and simple astigmatism. Presbyopia was defined as the need for near addition >= +1.00 D to achieve functional near vision at 37 cm (J1 level).

Uncorrected refractive error was defined as presenting visual acuity worse than 20/30 that improved to 20/30 or better after subjective refraction.

The primary outcome was the prevalence of refractive errors among Yanomami Indigenous individuals. Secondary outcomes included the distribution of refractive error types and the change in visual acuity following refractive correction.

### Statistical analysis

Continuous variables were summarised as means and standard deviations; categorical variables were expressed as absolute frequencies and percentages. Prevalence estimates were reported with stratification by sex, age group, and Yanomami subethnic subgroup. Age-standardised prevalence estimates were calculated by the direct method.

Comparisons between presenting and best-corrected visual acuity were performed using paired-sample Student’s t tests. Associations between refractive errors and demographic variables were evaluated using chi-square tests or Fisher’s exact tests, as appropriate. Odds ratios (ORs) with corresponding 95% confidence intervals (CIs) were estimated by binary logistic regression with age as a continuous predictor. All statistical tests were two-sided; a p value of less than 0.05 was considered statistically significant. Analyses were performed using Microsoft Excel, Statistica version 12 (TIBCO Software Inc.), and StatPlus (AnalystSoft). Given the convenience sampling approach, analyses were interpreted descriptively, emphasising epidemiological patterns rather than causal inference.

### Ethics approval

The study was approved by the Research Ethics Committee of the Federal University of Roraima and the Brazilian National Research Ethics Commission (Approval No. 6.479.368-CEP/CONEP). All procedures adhered to the tenets of the Declaration of Helsinki.

## Results

### Participant characteristics

A total of 158 Yanomami individuals aged 5 years or older were included in the analysis, representing 56.8% of the eligible population present at CASAI-Y during the study period. Participants belonged to four officially registered Yanomami subethnic subgroups: Yanomami (65.8%), Sanuma (24.0%), Xirixana (7.0%), and Xiriana (3.2%), and originated from multiple regions within the DSEI-Y. The final sample comprised 102 males (64.6%) and 56 females (35.4%), with a mean age of 32.4 years (SD 13.8). Demographic characteristics are presented in Table 1.

**Table 1.** Demographic characteristics of participants (N=158)

|  | Male (n=102) | Female (n=56) | Total (N=158) |
| --- | --- | --- | --- |
| Mean age (SD), years | 33.5 (13.7) | 30.4 (13.9) | 32.4 (13.8) |
| **Age group, n (%)** |  |  |  |
| 0-19 | 19 (18.6) | 13 (23.2) | 32 (20.3) |
| 20-39 | 40 (39.2) | 27 (48.2) | 67 (42.4) |
| 40-59 | 42 (41.2) | 15 (26.8) | 57 (36.1) |
| 60-80 | 1 (1.0) | 1 (1.8) | 2 (1.3) |
| **Subethnic subgroup, n (%)** |  |  |  |
| Yanomami | 62 (60.8) | 42 (75.0) | 104 (65.8) |
| Sanuma | 31 (30.4) | 7 (12.5) | 38 (24.1) |
| Xirixana | 5 (4.9) | 6 (10.7) | 11 (7.0) |
| Xiriana | 4 (3.9) | 1 (1.8) | 5 (3.2) |

*SD = standard deviation. Data are n (%) unless otherwise stated.*

### Prevalence and pattern of refractive errors

Among the 158 participants, strict emmetropia was observed in 107 individuals (67.7%). A clear age-related decline was evident, from 100% in children aged 5-9 years (11/11) to 95.2% in adolescents aged 10-19 years (20/21), 80.6% in adults aged 20-39 years (54/67), and 38.6% in those aged 40-59 years (22/57). No emmetropic individuals were identified among participants aged 60 years or older.

Myopia was present in 16.5% of participants (26/158); all cases were low myopia and no high myopia (SE <= -6.00 D) was identified. Notably, myopia was completely absent in children aged 5-9 years (0/11; 0.0%). Prevalence increased progressively with age, from 4.8% in adolescents aged 10-19 years to 17.9% in adults aged 20-39 years and 21.1% in those aged 40-59 years. Sex distribution was similar, with 16.7% in males and 16.1% in females.

Astigmatism was the most prevalent refractive condition, affecting 24.1% of participants (38/158), and was predominantly observed in combination with spherical refractive errors. Hyperopia was present in 13.3% (21/158), with all cases classified as mild. Among participants aged 35 years or older (n=75), presbyopia was identified in 39 individuals (52.0%), with a mean near addition of +2.04 D. Notably, 10 of the 39 presbyopic individuals (25.6%) were strict emmetropes who required near addition without distance correction. The full prevalence and classification of refractive errors are presented in Table 2.

**Table 2.** Prevalence and classification of refractive errors according to IMI criteria (N=158)

| Classification | n | % | Age-std % | OR/year (95% CI); p |
| --- | --- | --- | --- | --- |
| Emmetropia (SE -0.50 to +0.50 D) | 107 | 67.7 | 67.7 | 0.93 (0.91-0.95); <0.001 |
| Any refractive error | 51 | 32.3 | 36.1 | — |
| Myopia (SE ≤ -0.50 D) | 26 | 16.5 | 15.9 | 1.045 (1.010-1.081); 0.011 |
| Low myopia only | 26 | 16.5 | — | — |
| High myopia (SE ≤ -6.00 D) | 0 | 0.0 | — | — |
| Hyperopia (SE ≥ +0.50 D) | 21 | 13.3 | — | 1.205 (1.113-1.304); <0.001 |
| Mild hyperopia only | 21 | 13.3 | — | — |
| Astigmatism (cylinder ≥ 1.00 D) | 38 | 24.1 | 30.7 | 1.101 (1.059-1.144); <0.001 |
| Compound myopic astigmatism | 20 | 12.7 | — | — |
| Compound hyperopic astigmatism | 14 | 8.9 | — | — |
| Simple astigmatism | 5 | 3.2 | — | — |
| Presbyopia (age ≥35 years; n=75) | 39 | 52.0 | — | r=0.355; p=0.002 |
| Uncorrected RE with reduced VA | 40 | 25.3 | — | t=7.84; p<0.001 |

*SE = spherical equivalent; D = dioptre; OR = odds ratio; CI = confidence interval; VA = visual acuity; RE = refractive error. Age-standardised estimates by direct standardisation. Age ≥60 years: n=2, interpret with caution. Refractive categories not mutually exclusive except strict emmetropia. All cylinder values negative.*

### Age-related associations and regression analysis

Binary logistic regression confirmed significant age-related associations for all three principal refractive errors. The odds of myopia increased by 4.5% per year of age (OR 1.045; 95% CI 1.010-1.081; p = 0.011). Hyperopia showed the strongest age-related effect (OR 1.205; 95% CI 1.113-1.304; p < 0.001), largely reflecting its concentration in adults aged 40 years and older. Astigmatism also increased significantly with age (OR 1.101; 95% CI 1.059-1.144; p < 0.001). No significant associations were observed with sex or subethnic subgroup for any refractive error. Full regression results are presented in Table 3.

**Table 3.** Association between age and refractive errors: binary logistic regression (N=158)

| Refractive error | $\beta$ (SE) | OR per year | 95% CI | p value |
| --- | --- | --- | --- | --- |
| Myopia | 0.044 (0.017) | 1.045 | 1.010-1.081 | 0.011 |
| Hyperopia | 0.186 (0.041) | 1.205 | 1.113-1.304 | <0.001 |
| Astigmatism | 0.096 (0.020) | 1.101 | 1.059-1.144 | <0.001 |

*OR = odds ratio; CI = confidence interval; SE = standard error. All models include age as continuous predictor. p values from Wald statistics.*

### Uncorrected refractive error and visual acuity

A total of 40 participants (25.3%) presented with reduced distance visual acuity attributable to uncorrected refractive error. In 27 of these cases (67.5%), visual acuity improved significantly following optical correction, often reaching normal levels (mean improvement in decimal acuity: 0.32, SD 0.21; paired t-test: t = 7.84, df = 39, p < 0.001; Pearson’s r = 0.62, p < 0.001). The remaining 13 cases (32.5%) were potentially linked to non-refractive causes such as cataract or other ocular conditions.

Trends toward association were observed between pterygium (8.2%) and astigmatism (OR 2.22; 95% CI 0.60-8.18; p = 0.226), and between cataract (7.6%) and astigmatism (OR 3.57; 95% CI 0.92-13.86; p = 0.066), though neither reached statistical significance, likely owing to the small size of the affected subgroups. No association was observed between cataract and myopia (OR 0.68; 95% CI 0.08-5.75; p = 0.625). These findings are summarised in Table 4.

**Table 4.** Associations between comorbid ocular conditions and refractive errors.

| Condition (prevalence) | Outcome | OR (95% CI) | p value | Interpretation |
| --- | --- | --- | --- | --- |
| Pterygium (8.2%; n=13) | Astigmatism | 2.22 (0.60-8.18) | 0.226 | Trend; underpowered |
| Cataract (7.6%; n=12) | Astigmatism | 3.57 (0.92-13.86) | 0.066 | Borderline |
| Cataract (7.6%; n=12) | Myopia | 0.68 (0.08-5.75) | 0.625 | No association |

OR = odds ratio; CI = confidence interval. Fisher exact test used throughout. Analyses are hypothesis-generating.

## Discussion

Recent data from a 2025 study on Yanomami visual status reported a prevalence of moderate or worse visual impairment and blindness of 15.8% based on presenting visual acuity, decreasing to 8.2% with best-corrected visual acuity, with cataract and uncorrected refractive error as the leading causes.^1^□ In that analysis, uncorrected ametropias accounted for 74.3% of initial visual impairment, and significant visual acuity gains were observed after refractive correction. The present article provides the first detailed characterisation of the Yanomami refractive profile, expanding those findings to describe distribution by type, magnitude, age, sex, and subethnic subgroup, and evaluating associations with pterygium, cataract, and astigmatism.

The prevalence of strict emmetropia in the Yanomami population was high at 67.7%, consistent with a remote, low near-work environment. Yanomami emmetropia was higher than in Indigenous Brazilians from Avai/SP (approximately 45%),^1^□ rural South Indian adults aged 40 years or older (approximately 50%),^1^□ and substantially higher than in the Barbados Eye Study (approximately 31% in adults aged 40-84 years)^2^□ and very elderly European cohorts.□ Rural Southeast Asian populations showed comparable rates (approximately 65%).^1^□ These differences likely reflect contrasting degrees of urbanisation, age structure, and environmental exposure.

The Yanomami pattern of high emmetropia in younger age groups aligns with evidence for a protective role of sustained outdoor exposure and minimal near-work demands during childhood. Meta-analytic data demonstrate a dose-response relationship between time outdoors and reduced myopia incidence.^21^ Consistent with this, myopia was completely absent in children aged 5-9 years in the present study. However, intense lifelong solar exposure may have a dual effect: while high ambient light levels protect against developmental myopia through dopamine-mediated mechanisms,^21^ cumulative ultraviolet exposure may accelerate age-related lenticular changes,^22,23^ partially explaining the marked emmetropia decline observed after mid-life in this cohort.

The myopia prevalence of 16.5% is lower than in urban Brazilian adults (30-40%) but consistent with Amazonian Indigenous groups (10-18%) and rural Australian Aboriginal populations (11-15%),^11^ likely reflecting the protective effects of extensive outdoor activity and low near-work exposure. All cases were low myopia, and the complete absence of high myopia is a notable finding with potential implications for understanding the contribution of environmental factors to myopia severity globally.

Astigmatism at 24.1% mirrors patterns observed in tropical Indigenous populations, potentially reflecting corneal changes associated with chronic ultraviolet exposure. Hyperopia showed a strong age-related increase (OR 1.205 per year; p < 0.001), largely concentrated in adults aged 40 years and older, consistent with age-related lenticular shifts. Presbyopia affected 52.0% of participants aged 35 years or older, confirming the substantial unmet need for near vision correction in this population.

Trends toward associations between pterygium and astigmatism, and between cataract and astigmatism, did not reach statistical significance, likely due to the small size of the affected subgroups. These hypothesis-generating findings warrant investigation in larger samples and align with the 2025 study’s identification of cataract as a key cause of persistent visual impairment after refractive correction.^1^□

The findings have direct implications for eye health planning in the Yanomami territory. The high proportion of participants (25.3%) with reduced visual acuity due to uncorrected refractive error, and the substantial improvement observed with optical correction (67.5%; p < 0.001), underscore the potential impact of spectacle provision programmes at CASAI-Y and in remote communities. Culturally adapted assessment tools, such as the pictogram charts used in this study, and teleophthalmology services could address the logistical barriers inherent to this setting.

Several limitations should be acknowledged. The facility-based design at CASAI-Y may introduce some degree of selection bias, as the population comprises both patients referred for health care and accompanying family members. While the presence of patients may skew the sample toward individuals with greater healthcare needs, accompanying individuals are generally healthy community members, which partially mitigates this concern. Small subgroup sizes limited the power to detect associations with comorbid conditions. Cultural factors, including lower participation rates among women and older individuals, may have influenced the demographic composition of the sample. Future research should employ longitudinal designs, genetic analyses, and territory-wide approaches to disentangle environmental and hereditary contributions to refractive error in this population.

## Conclusions

This study provides the first characterisation of the refractive profile of the Yanomami people. The population demonstrates a distinct myopia pattern consistent with a lifestyle shielded from urban risk factors, with complete absence of myopia in children and no high myopia detected. Despite this, refractive correction remains effectively inaccessible, leaving preventable visual impairment unaddressed and reflecting a profound health inequity. Corrective lens provision and culturally adapted eye care services represent high-impact, scalable interventions for this underserved community.

## Data Availability

Due to the nature of this study involving Indigenous populations, individual participant data and associated documents cannot be shared without prior consultation and formal authorization from the communities involved. Any request for access to deidentified participant data and the data dictionary will require approval by the relevant Indigenous community leadership, submission and approval of a detailed research proposal, and signing of a Data Access Agreement ensuring respect for Indigenous data governance, confidentiality, and responsible data use. Data sharing requests may be addressed to the corresponding author and each request will be reviewed individually, subject to community approval and ethical oversight.

## Contributors

Maria Christina Chagas Ferreira: conceptualization, data curation, formal analysis, investigation, methodology, project administration, resources, writing (original draft and review and editing), and final responsibility for the decision to submit for publication.

Marcos Antonio Pellegrini: conceptualisation, methodology, resources, supervision.

Bianca Jorge Sequeira: formal analysis, methodology, supervision, writing (original draft and review and editing).

## Declaration of interests

All authors declare no conflicts of interest related to the content of this manuscript. No financial support, provision of study materials, medical writing assistance, or article processing charges were received for the development of this manuscript.

## Acknowledgements

We acknowledge the contributions of all individuals and institutions who supported the development of this study. We are especially grateful for the trust of the participants, whose collaboration was essential to the successful completion of this work.

## Declaration of generative AI and AI-assisted technologies in the writing process

During the preparation of this work the authors used ChatGPT (OpenAI, San Francisco, CA) to support the editing of text and improvement of language clarity in English. After using this tool, the authors carefully reviewed, verified, and edited all generated content and take full responsibility for the integrity and accuracy of the final version of the manuscript.

## Notes

**Declaration of competing interests:** All authors declare no conflicts of interest related to the content of this manuscript.

### Competing Interest Statement

The authors have declared no competing interest.

### Author Declarations

The Research Ethics Committee (CEP) of the Universidade Federal de Roraima (UFRR) and the National Research Ethics Commission (CONEP) of the Brazilian National Health Council gave ethical approval for this work (Approval No. 6.479.368-CEP/CONEP).

